# Mechanistic insights into heterogeneous radiofrequency ablation effects at the left atrial posterior wall during pulmonary vein isolation

**DOI:** 10.1101/19008706

**Authors:** David R. Tomlinson

## Abstract

**Background:** Independent investigations demonstrate greater radiofrequency (RF) ablation effects at left- sided left atrial posterior wall (LAPW) sites.

**Objective:** To investigate mechanisms underlying RF ablation heterogeneity during contact-force (CF) and VISITAG™ Module (Biosense Webster)-guided pulmonary vein isolation (PVI).

**Methods:** Consecutive patients undergoing PVI during atrial overdrive pacing comprised 2 cohorts: intermittent positive pressure ventilation (IPPV, 14-16/min, 6-8ml/kg); high frequency jet ventilation (HFJV, 150/min, Monsoon III, Acutronic). Temperature-controlled (17ml/min, 48°C) RF data was retrospectively assessed at first-annotated (target 15s) LAPW sites: 30W during IPPV; 20W at left-sided sites during HFJV.

**Results:** Twenty-five and 15 patients underwent PVI during IPPV and HFJV, respectively. During IPPV, left versus right-sided median impedance drop (ImpD) was 13.6Ω versus 9.9Ω (p<0.0001) respectively and mean time to pure R unipolar electrogram (UE) morphology change 4.9s versus 6.7s (p=0.007) respectively. During HFJV, ImpD was greater at left-sided sites (9.7Ω versus 7.4Ω, p=0.21) and time to pure R UE significantly shorter: 4.3s versus 6.1s (p=0.02). Minimum case impedance subtracted from pre-RF baseline impedance (BI) generated site-specific ΔBI. Left-sided sites demonstrated significantly greater ΔBI, correlating strongly with Ln(ImpD) – IPPV r=0.84 (0.65 – 0.93), HFJV r=0.77 (0.35 – 0.93).

At right-sided sites, ΔBI and Ln(ImpD) were without correlation during IPPV, but correlation was modest during HFJV (r=0.54, -0.007 – 0.84).

**Conclusions:** ΔBI may usefully indicate catheter-tissue contact surface area (SA). Consequently, greater left-sided LAPW RF effect may result from greater contact SA and in-phase catheter-tissue motion; HFJV may reduce right-sided out-of-phase catheter-tissue motion. Modifying RF delivery based on ΔBI may improve PVI safety and efficacy.

## Introduction

Pulmonary vein electrical isolation (PVI) is considered a prerequisite for atrial fibrillation (AF) ablation success.^1^ However, delivering permanently overlapping and transmural (TM) lesions without complications is a complex undertaking; on the one hand, non-TM lesions result in late pulmonary vein (PV) electrical gaps and recurrent AF^2^, whereas excessive focal radiofrequency (RF) energy delivery risks life-threatening extra-cardiac thermal trauma.^3^

Recently, RF annotation modules have been developed in an effort to provide guidance towards appropriate “per-site” RF delivery during RF PVI. These include novel systems for assessing the adequacy of “per-site” RF lesions, using weighted formulae incorporating catheter-tissue contact-force (CF), RF power and duration.^4^ Sites of RF application are automatically displayed on a 3-D electroanatomical map using objective descriptors of catheter stability, theoretically facilitating the development of safe, effective and reproducible PVI protocols.^5^ Subsequent studies have demonstrated how, following the derivation of putative ideal “per-site” targets for RF delivery via retrospective analyses of VISITAG™ Module annotation (Biosense Webster Inc., Diamond Bar, CA), >90% freedom from implantable loop recorder (ILR)-detected AF may result from active CF and VISITAG™ Module guidance during PVI.^6^ However, in this “CLOSE-guided” PVI 85 patient cohort, 75% experienced intra-oesophageal temperature rise >38.5°C (ITR, SensiTherm™, Abbott), totalling 183 episodes.^7^ Furthermore, the significantly greater proportion of ITR events occurring at the left-side of the LAPW indicates possible heterogeneity of RF effect at the LA posterior wall (LAPW). Such heterogeneity of RF effect during PVI has been demonstrated previously, with independently conducted investigations providing evidence towards significantly greater RF effect at left-sided left atrial sites.^8,9^ More recently and via retrospective analyses of VISITAG™ Module annotated sites, ∼30% greater RF effect was demonstrated at the left side of the LAPW, evidenced by significantly greater absolute impedance decrease and shorter time to pure R unipolar electrogram (UE) morphology change^10^ – a histologically validated marker of transmural (TM) RF effect in atrial tissue, *in vivo*.^11,12^ Importantly, off-line analyses of exported catheter RF and position data indicated that these findings were neither due to differences in RF duration, nor simply greater catheter instability and/or lower CF at right-sided LAPW sites.^10^

Taken together, these data indicate significantly greater RF effect at the left side of the LAPW of clinical relevance, yet without mechanistic explanation. One possibility is the occurrence of cardiac and/or respiratory cycle-induced site-specific differences in the magnitude of out-of-phase (i.e. sliding) catheter-tissue interaction. While it is neither desirable nor technically possible to perform PVI during complete atrial and ventricular standstill, high frequency jet ventilation (HFJV) provides means to eliminate respiratory cycle-induced changes in catheter-tissue interaction. Therefore, the aim of this present report was to retrospectively assess the magnitude of RF effects at first-ablated left and right-sided LAPW sites in consecutive cohorts of patients undergoing PVI during either intermittent positive pressure ventilation (IPPV), or HFJV.

## Methods

Single-operator (DRT) CF and VISITAG™ Module-guided PVI was performed employing a previously reported protocol^10^ in consecutive, unselected adult patients with symptomatic AF undergoing first-time PVI according to current treatment indications.^1^ All procedures were undertaken using general anaesthesia (GA) with endotracheal intubation. Single transseptal access was obtained with an SL1 (Abbott Cardiovascular, St Paul, MN) sheath, following which either a NaviStar^®^ THERMOCOOL^®^ SMARTTOUCH™ (ST) F curve or NaviStar^®^ EZSTEER^®^ THERMOCOOL^®^ SMARTTOUCH™ D/F catheter (Biosense Webster) via an Agilis™ NxT sheath (Abbott) was placed in the LA via the first transseptal site. ACCURESP™ respiratory training was undertaken pre-ablation and applied as required to the CARTO^®^ geometry, created using a LASSO^®^ Nav catheter (Biosense Webster). Where respiratory motion was insufficient to trigger the ACCURESP™ detection threshold the tidal volume was never deliberately increased, but where ACCURESP™ respiratory adjustment threshold was exceeded, VISITAG™ Module filter preferences during ablation employed ACCURESP™ set “off” to avoid RF annotation error.^13^ The IPPV cohort employed fixed 6-8ml/kg ventilation at14-16 breaths per minute, guided by end-tidal CO_2_; positive end-expiratory pressure 5cmH_2_O; 50% inspired oxygen concentration. For the HFJV cohort, GA was induced and maintained with a total intravenous anaesthesia technique guided by depth of anaesthesia monitoring (BIS™, Medtronic Inc., St Paul, MN). IPPV was employed until the mapping and ablation phases of the procedure, whereupon HFJV (Monsoon III, Acutronic Medical Systems AG, Hirzel, CH) was used via a jet ventilation catheter; inspired oxygen concentration 60% (titrated to maintain oxygen saturations ≥95%); ventilation frequency 150 jets/min; driving pressure 1.0 bar with 1:1 inspiration to expiration ratio.

A SkinTact® RO01B30 indifferent electrode (Leonhard Lang GmbH, Innsbruck, AT) was placed on the shaved skin of the thigh posterolaterally, contralateral to any orthopaedic prostheses. VISITAG™ Module and CF-guided PVI was performed during proximal pole CS pacing at 600ms using temperature-controlled RF (48°C, 17ml/min irrigation) via an EP Shuttle^®^ (Stockert GmbH, Freiburg, DE) generator and ThermoCool® SmartTouch® catheter using Agilis™ NxT sheath (Abbott) support. VISITAG™ Module filter preferences for automated RF annotation were: Positional stability range 2mm, tag display duration 3s; force-over-time 100% minimum 1g (the latter to ensure on-going RF annotation only in the presence of constant catheter-tissue contact^14^). Lesion placement was guided by VISITAG™ Module annotation, with the preferred site of first RF application at the LAPW opposite each superior PV ∼1cm from the PV ostium; in cases where constant catheter-tissue contact could only be achieved with maximal CF ≥70g, an adjacent LAPW site with lower peak CF was chosen. The target annotated RF duration at each first-ablated LAPW site was 15s, whereas ∼9-11s was the target for consecutive, adjacent sites during continuous RF delivery. Target inter-tag distance (ITD) ≤6mm was achieved using rapid movement of the catheter tip initiated via the Agilis sheath, aided by the distance measurement tool; point-by-point (PbP) RF was also applied as necessary to achieve this end-point.

For the IPPV cohort, 30W RF was used at all sites. However, in view of previous investigations demonstrating ∼30% greater RF effect at the left-side of the LAPW^10^, 20W was the peak power used during first encirclement at left-sided LAPW sites during HFJV. In addition to the RF duration targets described, annotated “per site” LAPW targets during HFJV included a minimum impedance drop of 3Ω. Following completion of circumferential PVI (entrance and exit block), spontaneous recovery of PV conduction was assessed and eliminated during a minimum 20-minute wait; dormant recovery was evaluated and eliminated a minimum of 20 minutes after the last RF. Neither oesophageal luminal temperature monitoring nor post-ablation endoscopic evaluation was employed.

This work received IRB approval for publication as a retrospective service evaluation. All patients provided written, informed consent.

### Analyses

Annotated first-site RF duration, mean CF and impedance drop (ImpD) data were obtained via the proprietary VISITAG™ Module export function. Bipolar electrogram (BE) peak-to-peak amplitude and the timing of unipolar electrogram (UE) morphology change from RS to pure R data were retrospectively obtained from CARTOREPLAY™ (Biosense Webster) as previously described.^10^ Exported text files were converted to Excel data and imported to GraphPad Prism version 4.03 (GraphPad Software, San Diego, CA). For correlation analyses, ImpD data was Ln transformed to achieve a normal distribution. Considering the known importance of the surface area (SA) of catheter-tissue contact towards RF lesion creation^15^, a hypothetical indicator of the ablation catheter-tissue SA at RF onset was retrospectively derived from VISITAG™ Module-based ablation catheter (unipolar) impedance data. This was calculated by subtracting the minimum case impedance value (from the exported “AblationData” file) from the immediate pre-RF onset impedance (i.e. “Base impedance”, BI), to derive the site-specific delta base impedance (ΔBI).

### Statistics

Comparisons were made based on the means and standard deviations [SD] of biophysical data (i.e. ILD, RF duration, CF, ImpD), or medians along with the 1^st^ and 3^rd^ quartile (IQR), where these data were determined to be skewed. Differences in the biophysical data between IPPV and HFJV, and left and right-sided LAPW sites were tested using the unpaired t-test, or the Mann-Whitney test where data could not be assumed to be normally distributed. The Pearson or Spearman correlation coefficient was calculated to determine the strength of association between Ln (ImpD) and both BI and ΔBI, as appropriate. In this exploratory analysis, significance was set at the 5% level.

## Results

The IPPV cohort comprised 25 patients (November 2016 – May 2017): 13 persistent AF, 12 PAF; 19 male (76%); mean age 57 [14] years and CHA_2_DS_2_-VASc score 1.3 [1.3]. The HFJV cohort comprised 15 patients (July 2018 – June 2019): 9 persistent AF, 6 PAF; 12 male (80%); mean age 60 [9] years and CHA_2_DS_2_-VASc score 1.2 [1.1]. Complete PVI with the elimination of spontaneous and/or dormant recovery of PV conduction was achieved following 16.2 [3.1] and 20.0 [4.6] minutes of RF for IPPV and HFJV cohorts, respectively. For the IPPV cohort, pulmonary vein (PV) carina RF was applied in 18/25 patients (72%); i.e. left PV carina in 8/25 (32%) and right PV carina in 16/25 (64%). For the HFJV cohort, PV carina RF was applied in 13/15 patients (87%); i.e. left PV carina in 10/15 (67%), right PV carina only 10/15 (67%). There were no procedural complications.

In the IPPV cohort, 3 first-ablated sites were excluded from analysis; inadvertent catheter displacement at two left-sided sites resulted in RF annotation termination at 4.4s and 7.9s, while catheter displacement at RF onset resulted in 5.0s non-annotated RF delivery for one right-sided site. In the HFJV cohort, 4 first-ablated sites were excluded from analysis: For one left-sided site, DC cardioversion (DCCV)-refractory AF was present, while inadvertent catheter displacement resulted in RF annotation termination at 7.1s in another; pure R UE morphology change was not achieved at one left-sided site with pre-ablation bipolar electrogram (BE) amplitude 0.7mV, annotated RF duration 14.7s, mean CF 21g, and ImpD 8.6Ω - excepting the one case of DCCV-refractory AF, this was the lowest pre-ablation BE amplitude in the HFJV cohort and was considered an outlier; inadvertent catheter displacement resulted in RF annotation termination at 7.6s for one right-sided site.

### Biophysical data: IPPV cohort

First-ablated LAPW site biophysical data according to ventilatory protocol and PV pair proximity at the LAPW are shown in table 1. During IPPV and using 30W, annotated RF duration was in keeping with the protocol; i.e. median 14.9s versus 15.0s, left PV (LPV) versus right PV (RPV) aspects of the LAPW respectively. There was no significant difference in pre-ablation BE amplitude; median 1.5mV versus 1.7mV (p=0.95), LPV and RPV respectively. Mean CF was significantly greater at RPV LAPW sites; i.e. 16.5g versus 11.2g (p=0.003), RPV and LPV respectively. During IPPV, both ImpD and time to pure R UE morphology change data indicated significantly greater RF effects at the left side of the LAPW: median ImpD 13.6Ω versus 9.9Ω (p<0.0001), LPV versus RPV respectively; mean time to pure R UE morphology change 4.9s versus 6.7s (p=0.007), LPV versus RPV respectively.

**Table 1:** Biophysical data according to ventilatory protocol and pulmonary vein (PV) pair proximity (i.e. left and right PV, LPV and RPV, respectively) at first-ablated LAPW sites. Data shown are mean [SD] and median (1^st^ – 3^rd^ quartile), as appropriate (BE, CARTOREPLAY™-derived peak-to-peak bipolar electrogram amplitude; BI, baseline impedance; ΔBI, minimum case impedance subtracted from BI; Ln(ImpD), natural log of the annotated impedance drop).

|  | IPPV (N=25) |  |  | HFJV (N=15) |  |  |
| --- | --- | --- | --- | --- | --- | --- |
|  | LPV (30W; N=23) | RPV (30W; N=24) | p | LPV (20W; N=12) | RPV (30W; N=14) | p |
| RF duration (s) | 14.9<br>(14.3 – 15.7) | 15.0<br>(14.6 – 15.4) | 0.70 | 14.5<br>(14.1 – 15.1) | 14.5<br>(13.7 – 15.0) | 0.40 |
| Pre-RF BE (mV) | 1.5<br>(1.1 – 3.1) | 1.7<br>(1.0 – 2.3) | 0.95 | 1.5<br>(1.1 – 2.9) | 1.7<br>(1.0 – 3.1) | 0.94 |
| Mean CF (g) | 11.2<br>(9.0 – 13.4) | 16.5<br>(12.2 – 18.4) | 0.003 | 12.5<br>(10.0 – 14.9) | 12.0<br>(9.4 – 12.2) | 0.61 |
| Impedance drop ($\Omega$) | 13.6<br>(12.2 – 24.4) | 9.9<br>(7.9 – 11.6) | <0.0001 | 9.7<br>(7.2 – 13.7) | 7.4<br>(6.7 – 9.6) | 0.21 |
| Time to pure R UE (s) | 4.9<br>[2.1] | 6.7<br>[2.5] | 0.007 | 4.3<br>[1.4] | 6.1<br>[2.1] | 0.02 |
| BI ($\Omega$) | 182<br>(177 – 196) | 175<br>(170 – 180) | 0.002 | 183<br>(175 – 201) | 174<br>(160 – 182) | 0.03 |
| ΔBI ($\Omega$) | 45.0<br>[15.2] | 35.3<br>[9.0] | 0.01 | 45.7<br>[9.4] | 32.4<br>[9.0] | 0.004 |
| Pearson/Spearman r: BI vs Ln (ImpD) | 0.66<br>(0.34 – 0.84) | 0.33<br>(-0.10 – 0.65) | NA | 0.59<br>(0.03 – 0.87) | 0.33<br>(-0.26 – 0.74) | NA |
| Pearson/Spearman r: ΔBI vs Ln (ImpD) | 0.84<br>(0.65 – 0.93) | 0.26<br>(-0.18 – 0.61) | NA | 0.77<br>(0.35 – 0.93) | 0.54<br>(-0.007 – 0.84) | NA |

During IPPV the mean BI was significantly greater at left-sided LAPW sites; i.e. median 182Ω versus 175Ω (p=0.002), LPV versus RPV. The mean minimum (total case) impedance was 141Ω, therefore the mean ΔBI was significantly greater at LPV versus RPV sites; i.e. 45.0Ω versus 35.3Ω (p=0.01), LPV versus RPV respectively. For left-sided LAPW sites, both BI and ΔBI demonstrated significant positive correlation with Ln(ImpD), but correlation was strongest for ΔBI: i.e. BI with Ln(ImpD) Pearson r=0.66 (0.34 – 0.84, p=0.0006); ΔBI with Ln(ImpD) r=0.84 (0.65 – 0.93, p<0.0001). For right-sided LAPW sites, neither BI nor ΔBI demonstrated significant correlation with Ln(ImpD); i.e. Spearman r=0.33 (-0.10 – 0.65, p=0.12) and r=0.26 (-0.18 – 0.61, p=0.23) for BI with Ln(ImpD), and ΔBI with Ln(ImpD) respectively (figure 1).

**Figure 1:**
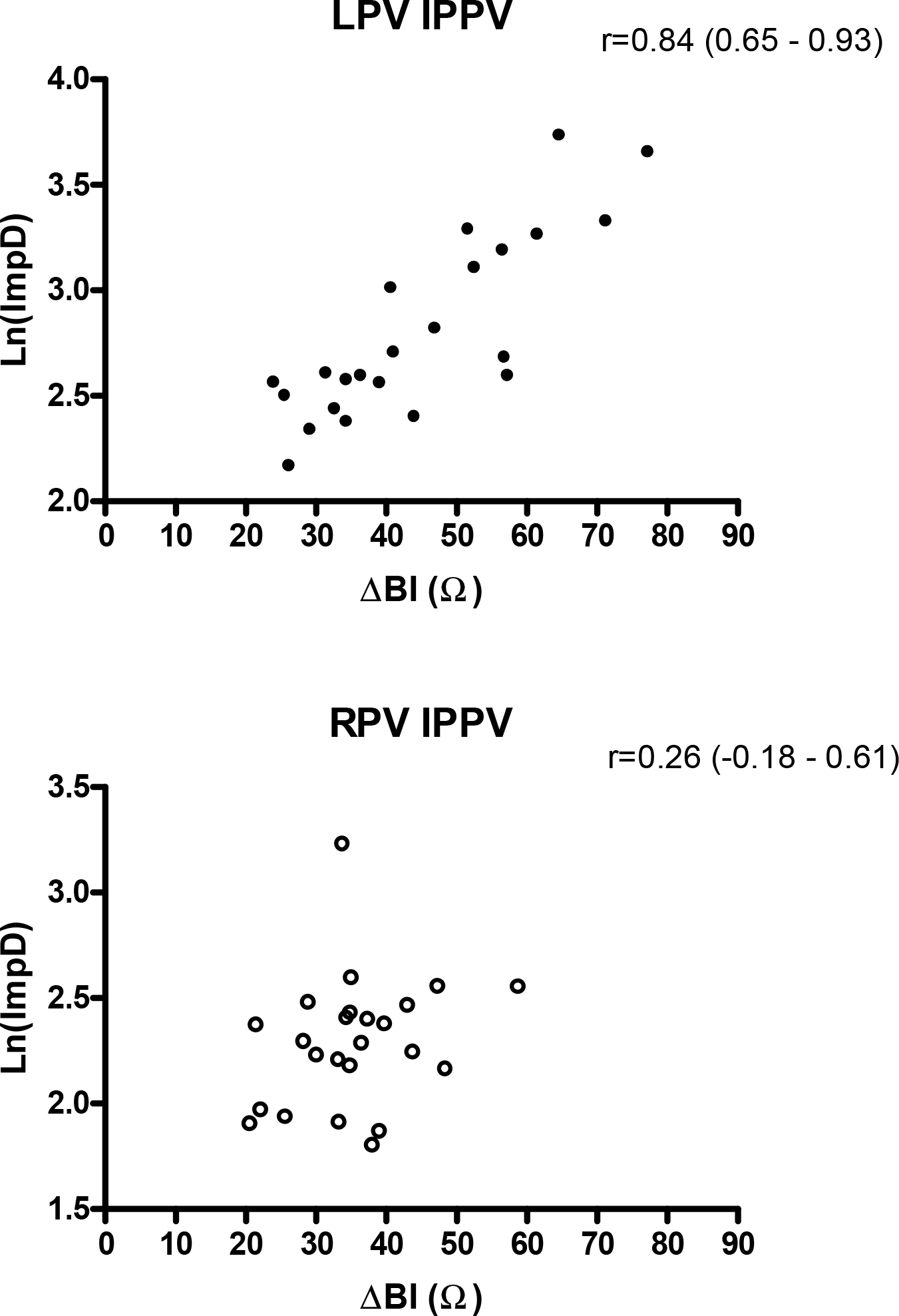
Correlation between ΔBI and Ln(ImpD) during IPPV according to PV proximity (Pearson / Spearman correlation is displayed, as appropriate).

### Biophysical data: HFJV cohort

During HFJV, and using 20W and 30W at left and right-sided LAPW sites respectively, annotated RF duration was in keeping with the protocol; i.e. median 14.5s, both PVs. There was no significant difference in pre-ablation BE amplitude; median 1.5mV versus 1.7mV (p=0.94), LPV and RPV respectively. There was no difference in mean CF during HFJV; i.e. 12.5g and 12.0g (p=0.61), LPV versus RPV respectively. During HFJV, median ImpD was greater at LPV sites (9.7Ω versus 7.4Ω, p=0.21) but this did not reach statistical significance. However, time to pure R UE morphology change was significantly shorter at LPV sites; i.e. mean 4.3s versus 6.1s (p=0.02), LPV versus RPV respectively.

During HFJV the mean BI was significantly greater at left-sided LAPW sites; i.e. 183Ω versus 174Ω (p=0.03), LPV versus RPV respectively. The mean minimum (total case) impedance was 143Ω and 136Ω, LPV and RPV respectively (p=0.25). Therefore, the mean ΔBI was significantly greater at LPV versus RPV sites; i.e. 45.7Ω versus 32.4Ω (p=0.004), LPV versus RPV respectively. At left-sided LAPW sites during HFJV both BI and ΔBI demonstrated significant positive correlation with Ln(ImpD), but correlation was strongest for ΔBI; BI with Ln(ImpD), Pearson r=0.59 (0.03 – 0.87, p=0.04), whereas ΔBI with Ln (ImpD), r=0.77 (0.35 – 0.93, p=0.004). At right-sided LAPW sites during HFJV there was no significant correlation between BI and Ln(ImpD); Spearman r=0.33 (-0.26 – 0.74, p=0.25). However, right-sided LAPW sites during HFJV demonstrated moderate positive correlation for ΔBI with Ln(ImpD); Spearman r=0.54 (-0.007 – 0.84, p=0.047, figure 2).

**Figure 2:**
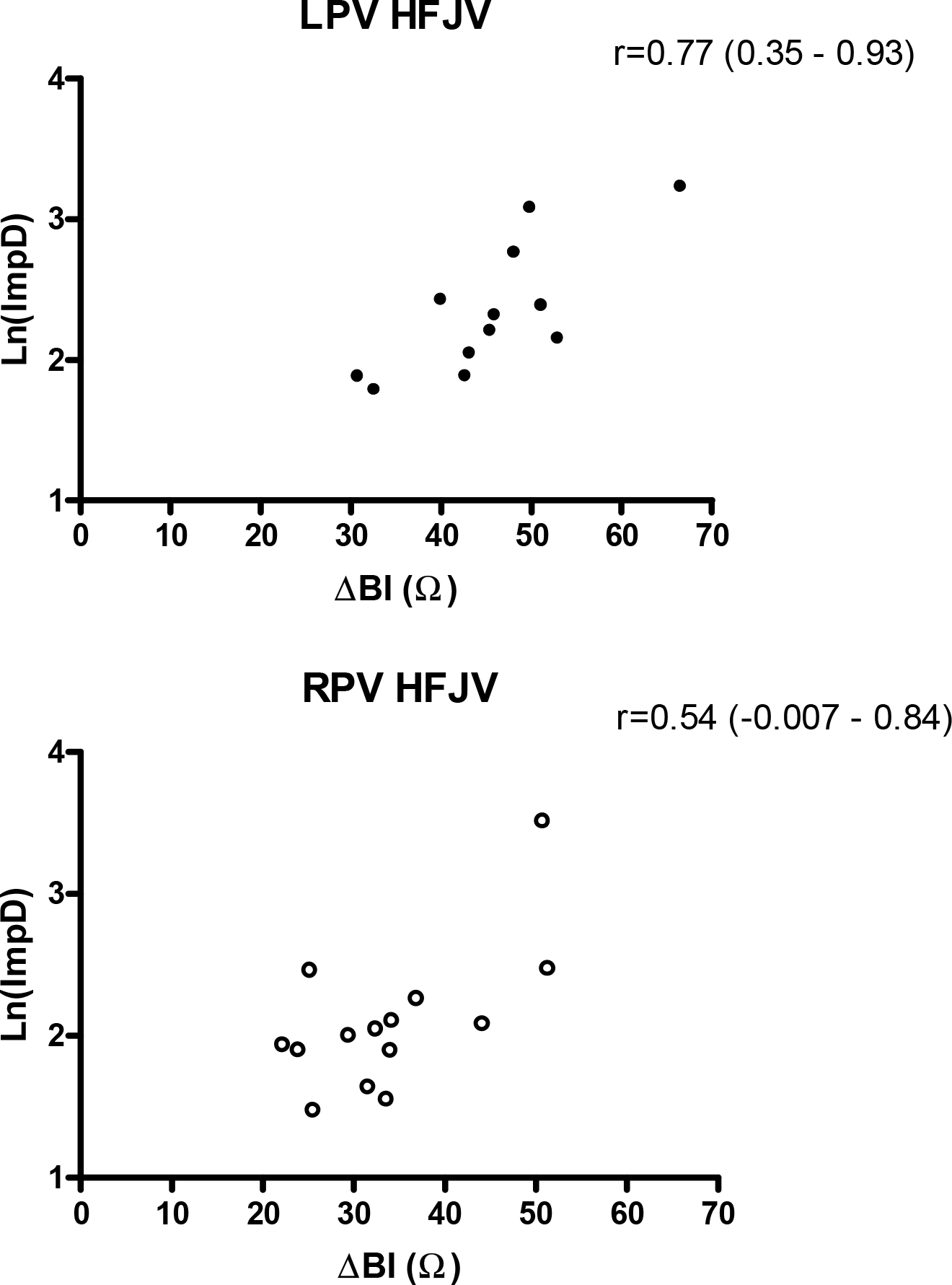
Correlation between ΔBI and Ln(ImpD) during HFJV according to PV proximity (Pearson / Spearman correlation is displayed, as appropriate).

There was no significant correlation between mean CF and Ln(ImpD) within any ventilatory group; i.e. r=-0.24, 0.07, 0.02 and -0.10 for LPV IPPV, RPV IPPV, LPV HFJV and RPV HFJV, respectively.

## Discussion

The main finding of this present report is that heterogeneity of RF effect at the LAPW was not eliminated using a HFJV protocol; i.e. greater effect remained evident at left-sided sites, despite reducing left-sided LAPW power delivery by 30%. Therefore, although heterogeneity of RF effect cannot be completely explained on the basis of respiratory motion-induced differences catheter-tissue interaction, these present data can be used to derive hypotheses towards other possible mechanisms underlying heterogeneity of RF effects during PVI.

### Hypothetical mechanisms underlying heterogeneity of RF effects *in vivo*

In this present experimental set-up, identical and objectively annotated parameters for position and CF stability – the two presently measureable components of catheter “stability” – were prospectively employed in order to meet a suitable definition of a “stable” point: (1) Position stability range 2mm with ACCURESP “off” – VISITAG™ Module logic previously shown to satisfy “stable point” criteria^13^, and; (2) force-over-time 100%, minimum 1g CF – self-evidently, constant catheter-tissue contact is a prerequisite at “stable” sites.^14^ Previous investigations have demonstrated that greater RF effect at left-sided LAPW sites was not simply due to measurably superior position stability.^10^ Therefore, assuming equal RF generator power delivery, greater RF effect in this *in vivo* model must result from either greater catheter-tissue interaction stability as a result of significantly lower *out-of-phase* (i.e. sliding) catheter-tissue interaction, and/or a greater SA of catheter-tissue contact.

From a theoretical standpoint, if an ablation catheter is kept “positionally stable” and (knowingly) remains stable throughout “per-site” RF delivery without significant out-of-phase catheter-tissue interaction, differences in the SA of catheter-tissue contact will be the principal determinant of measureable differences of RF effects, since:

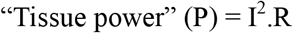

(I is current density and R, tissue resistance).^15^

In this theoretical model of stable catheter-tissue interaction, any suitable measure of the SA of catheter-tissue contact will demonstrate strong positive correlation with RF effects. Therefore, this present report’s finding of a strong positive correlation between ΔBI and Ln(ImpD) at left-sided LAPW sites indicates not only that ΔBI may represent a suitable indicator of the SA of catheter-tissue contact *in vivo*, but also that catheter-tissue interaction at left-sided LAPW sites is predominantly *in-phase*. At the right-sided LAPW sites, the significantly lower ΔBI supports the proposition that ΔBI provides a suitable measure of catheter-tissue contact SA. However, during IPPV and despite ablation catheter motion remaining within identical pre-defined limits, the lack of significant correlation between ΔBI and Ln(ImpD) indicates significant and presently un-measureable, *out-of-phase* catheter-tissue interaction. In contrast, the finding of a modest positive correlation between ΔBI and Ln(ImpD) at right-sided LAPW sites during HFJV indicates that the operative environment during HFJV is characterised by a reduction in out-of-phase catheter-tissue interaction, but not to the same level of in-phase stability as that inferred using these methods at left-sided LAPW sites.

Possible mechanisms for significant differences in the SA of catheter-tissue contact comparing left and right-sided sites could include either/or both of: (1) Differences in the angle of the catheter tip with respect to the target tissue, such that at some sites the contact surface may include both the (tip) face and lateral aspects of the catheter tip, whereas at others, contact is limited to the surface of the (tip) face only; (2) Differences in tissue compliance, with greater SA resulting from more compliant tissue enveloping the catheter tip, potentially even at lower CF. These present data do not permit further conclusions to be drawn regarding the relative importance of these possible mechanisms *in vivo*, however, greater impedance drop has been demonstrated during PVI with parallel or oblique catheter-tissue angle (30° – 145°) compared to perpendicular (0 – 30°).^8^

### Pre-ablation impedance as a possible determinant of RF effect *in vivo*

Previous investigations during slow pathway and accessory pathway RF ablation demonstrated strong positive correlation between the initial generator impedance (GI) and electrode temperature during RF delivery.^16^ In a pig thigh model of RF ablation using a linear multi-electrode catheter, greater baseline (unipolar) impedance was demonstrated with greater applied downward force, and there was a strong positive correlation between baseline impedance and lesion depth and width.^17^ More recently and using an ablation catheter with mini-electrodes incorporated in the distal electrode (Rhythmia™ and IntellaNav MiFi™ OI, Boston Scientific, Marlborough, MA), the predictive utility of baseline generator and local impedances (LI – taken as the maximal value from all three mini electrodes) were investigated during PVI. The baseline GI demonstrated weak positive correlation with subsequent RF-induced GI drop (adjusted R^2^ = 0.06, p<0.001). However, higher baseline LI predicted higher LI drop during ablation (adjusted R^2^ = 0.41, p<0.001).^18^ Lastly, finite element modelling of a 7F 4mm ablation catheter and homogeneous tissue slab (4mm thickness) with simulations of varying depth and angle of catheter-tissue interaction, demonstrated a very strong positive correlation between increasing contact area and impedance (R=0.97).

Moreover, while the impedance increased at greater depth of catheter-tissue penetration, the catheter angle was a significant determinant of impedance; incrementally greater impedance was noted as the angle was decreased from 90° to 15° in 15° steps. Importantly, catheter angle was a more important determinant of impedance at greater modelled depth of tissue penetration.^19^

### Clinical implications

The inference of both significantly greater *in-phase* catheter-tissue interaction and greater SA of stable contact at left-sided LAPW sites provides the perfect milieu for inadvertent extra-cardiac thermal trauma from a “one-size-fits-all”, single RF power and duration delivery protocol during PVI. Similarly, ablation protocols employing identical LAPW RF targets using theoretical composite measures of RF delivery (i.e. Ablation Index – AI, or VISITAG SURPOINT™, Biosense Webster – and lesion size index – LSI, Abbott) will also result in heterogeneous tissue injury during PVI, perpetuating this risk.^4,6^ This is particularly concerning in view of the more frequently occurring left-sided oesophageal position^20^ and high case fatality rates of ablation-induced atrio-oesophageal fistula.^21^ Therefore for greater safety, operators may wish to consider adopting protocols involving lower RF power, shorter RF duration and/or lower AI/LSI targets at the left side of the LAPW, or simply utilising more direct and *in vivo* validated measures of tissue RF effect including impedance drop and/or pure R UE morphology change.^10,22–24^

Since ΔBI may represent a useful site-specific *pre-ablation predictor* of RF effect, operators may wish to mimic this described operative “stability set-up”. Specifically, with ΔBI representing a putative predictor of RF effect within this reproducible “operative stability environment”, the risk of extra-cardiac thermal trauma during RF delivery using higher power (50-90W) over very short ablation intervals (e.g. 3-5s)^25,26^ – i.e. within potentially very narrow therapeutic windows and involving significant latency^27^ – may be reduced by modified protocols incorporating ΔBI data. In contrast, protocols using deep conscious sedation with consequent greater random variability of respiratory profile and occasional inadvertent patient movement are unlikely to demonstrate similar predictive utility of ΔBI towards subsequent “per-site” RF effects.

### Limitations

Time to pure R UE morphology change is an imperfect indicator of RF effect in the absence of left atrial wall thickness data; i.e. shorter time to pure R alone does not necessarily indicate greater RF effect. The methodology for defining the site-specific ablation catheter-to-tissue contact SA described in this present report (i.e. ΔBI) is theoretical. Indeed, more appropriate methodology towards deriving putative “predictive” ΔBI values would be to subtract the pre-ablation “blood-pool” impedance from site-specific BI. However, at the outset of these present investigations “blood-pool” impedance data were not known to be of potential use, and there were no means to obtain these data retrospectively. Clearly, the hypothesis that pre-ablation ΔBI represents a site-specific predictor of RF effect during PVI requires further investigations using “blood-pool” impedance values.

These data are limited by their single operator status, applicability to the operative stability conditions described and choice of RF annotation logic using the VISITAG™ Module. However, such stringent catheter stability criteria alongside measureable (and visible to any operator) VISITAG™ Module data outputs may permit these present investigations to form the basis of knowingly reproducible RF delivery *in vivo*, both on an intra and inter-operator basis. These data should not be considered applicable to TactiCath™-based ablation (Abbott), since according to the dimensions of the ablation catheter tip electrode, angle-dependent changes in the SA of contact will result in catheter-specific differences in tissue RF current density and ablation effect.^28^

RF power output from the EP Shuttle^®^ RF generator has been shown to vary according to the pre-ablation impedance; i.e. comparing 30W at 150Ω versus 200Ω, power output at 200Ω was 14% greater, measuring 34.1W.^29^ However, differences between left and right-sided LAPW BI values in this present report (IQR 177 – 196 and 170 – 180 for LPV and RPV, respectively) were probably insufficient to importantly influence generator RF power output and furthermore, greater RF effect remained at left-sided sites during HFJV despite 30% lower power delivery at left-sided sites.

## Conclusions

These data support the hypothesis that greater RF ablation effect at left-sided LAPW sites results from significantly greater SA of catheter-to-tissue contact and in-phase catheter-tissue motion. However, HFJV may usefully reduce out-of-phase catheter-tissue motion at right-sided LAPW sites. Using suitably stable and reproducible conditions of catheter-tissue contact, ΔBI may represent a useful site-specific predictor of RF ablation effect, towards improving the safety and efficacy of PVI.

## Data Availability

No links to raw data provided, although original data files will be shared following any reasonable request.

## Acknowledgements

I am grateful to Cherith Wood, Daniel Newcomb and Ian Lines, Cardiac Physiologists, for their technical support into all cases conducted during this report and Dr David Adams and Dr Kate Holmes for establishing and delivering the HFJV protocol. I am also grateful to Robert Pearce and Vicky Healey (Biosense Webster Inc.) for additional technical assistance and to Noam Seker-Gafni, Tal Bar-on, Einav Geffen, Assaf Rubissa and colleagues at the Haifa Technology Center, Israel for their help with VISITAG™ Module technical queries. I am grateful to Katie Biscombe, John True, Joanne Hosking and Adam Streeter (Department of Medical Statistics, Plymouth University Peninsula Schools of Medicine and Dentistry) for their previous R code development and analyses permitting reference to previously identified measures of catheter stability in this present report.

